# Effectiveness of nirsevimab against hospitalization for RSV bronchiolitis according to RSV lineages

**DOI:** 10.64898/2026.09.28.26364231

**Authors:** Mathieu Barbier, Corinne Levy, Robert Cohen, Zaba Valtuille, Zein Assad, Jee-seon Yang, Inès Fafi, Léa Lenglart, Stéphane Béchet, Lindsay Osei, Elise Launay, Christèle Gras-Le Guen, Vincent Gajdos, Eric Jeziorski, Loïc de Pontual, Romain Basmaci, Marie-Anne Rameix-Welti, Naim Ouldali, Slim Fourati

## Abstract

**Importance:** Nirsevimab, a monoclonal antibody against RSV, was implemented in France in September 2023. Recently, rare resistance-associated substitutions were described in RSV but the effectiveness according to lineages remains unclear.

**Objective:** We aimed to estimate the effectiveness of nirsevimab against hospitalization for RSV bronchiolitis according to RSV lineages.

**Design:** We conducted a multicenter test-negative case-control study from October 15, 2023 to March 15, 2025.

**Setting:** This study was based on data from three national multicentric prospective studies. The 12 participating centers were secondary or tertiary hospital with a pediatric department located in France.

**Participants:** We included all children younger than 12 months who were hospitalized for RSV bronchiolitis in one of the participating centers in France during the two first RSV epidemic seasons following the implementation of nirsevimab (from October 15, 2023 to February 29, 2024, and from October 10, 2024 to March 15, 2025).

**Main outcomes and measures:** The main outcome of the study was the RSV test result of bronchiolitis cases. RSV whole-genome sequencing was performed using a tiled amplicon approach covering the complete viral genome, as previously described in the POLYRES studies. We estimated the effectiveness of nirsevimab by a multivariable logistic regression model adjusted for potential confounders. We estimated the adjusted risk of immunization failure for each lineage by calculating relative odds ratio to a reference lineage. A range of sensitivity analyses were conducted.

**Results:** We included 3019 infants with bronchiolitis among which 2273 had a positive RSV test (case patients) and 746 had a negative RSV test (control patients). Overall, 373/2273 (16.4%) case patients were immunized by nirsevimab versus 423/746 (56.7%) control patients. The adjusted effectiveness of nirsevimab against hospitalization for bronchiolitis was 88.4% (95% CI, 84.5–91.4) with RSV-B and 79.4% (95% CI, 73.7–83.8) with RSV-A. The risk of immunization failure was 3.2-fold (95% CI 1.8–5.7) higher for lineages derived from A.D.3 than for lineages derived from B.D.4.1.

**Conclusion and relevance:** In this multicentric case-control study, we observed that nirsevimab effectiveness was variable across RSV lineages, with a lower effectiveness against A.D.3 lineages compare to others. Further surveillance is needed to confirm these findings and assess their implication on RSV lineages epidemiology as nirsevimab use expands.

**KEYPOINT:** *Question:* Does the effectiveness of nirsevimab against hospitalization for RSV-bronchiolitis change based on the RSV lineages?

*Findings:* In this case-control test negative design study that include 3019 infants, the effectiveness of nirsevimab against hospitalization for bronchiolitis was significantly higher with RSV-B than with RSV-A. Specifically, we found a 3.2-fold risk of immunization failure with lineages derived from A.D.3 than with lineages with highest effectiveness.

*Meaning:* This study suggest that nirsevimab effectiveness varies substantially across RSV lineages, with lineages derived from A.D.3 less susceptible to nirsevimab. This highlight the importance of molecular and clinical surveillance over the coming years of wide nirsevimab use.

## INTRODUCTION

Bronchiolitis is the most common lower respiratory tract infection in infants.^1^ Respiratory syncytial virus (RSV) is the main pathogen of bronchiolitis, with around 30 million cases, 3 million hospitalizations and up to 200, 000 deaths in children younger than five years worldwide each year.^2^ RSV is an enveloped RNA virus that belongs to the family *Pneumoviridae*. In humans, two groups, RSV-A and RSV-B co-circulate worldwide each year, and multiple lineages have been described within each group.^3^ Until 2023, palivizumab was the only preventive treatment approved for RSV, indicated for the high-risk children.^4^ Recently, nirsevimab, a long-lasting monoclonal antibody that binds the RSV prefusion F protein has been developed.^5^ Two multicenter randomized trials have shown the efficacy of nirsevimab in reducing medically attended RSV-associated lower respiratory tract infections in healthy term and pre-term infants.^6,7^ In France, nirsevimab became available in September 2023 for all infants during their first epidemic season of RSV. Several studies have demonstrated a high effectiveness of nirsevimab against RSV-related hospitalization and medically attended in primary care settings.^8–12^ The widespread implementation of nirsevimab has the potential to result in viral escape, thereby raising the risk of the spread of RSV strains resistant to nirsevimab. A recent multicenter French study analysed RSV genome sequencing of breakthrough infections and found resistance-associated substitutions in around 10% of RSV-B samples.^13,14^ While resistance-associated substitutions appear rare, the potential impact of RSV genetic diversity, across subtypes and lineages on nirsevimab effectiveness remains unknown. Because monoclonal antibodies recognize specific conformational epitopes, natural RSV genetic diversity across subtypes and lineages may lead to differences in susceptibility to neutralization even in the absence of established resistance-associated substitutions. In this context, we aimed to estimate the effectiveness of nirsevimab against hospitalizations for RSV-related bronchiolitis according to subtypes and lineages.

## 1. METHODS

### 1.1. Study design and participants

We conducted a national multicentric prospective case-control test-negative design study from October 15, 2023 to March 15, 2025, covering the first (2023–2024) and second (2024–2025) RSV seasons following the implementation of nirsevimab immunization. For each season, the inclusion period was defined by the RSV epidemic period in France, weekly monitored by *Santé Publique France*.^15^ The first inclusion period was between October 15, 2023 and February 29, 2024. The second inclusion period was between October 10, 2024 and March 15, 2025. The test-negative design, which is used to assess vaccine effectiveness and minimize confounding bias due to healthcare seeking behavior, has seen increased application in the estimation of RSV immunization program effectiveness.^16,17^

We included all children younger than 12 months who were hospitalized at one of the 12 participating centers for acute viral bronchiolitis during the study period and had an RSV diagnostic test at the admission or during their stay. Acute viral bronchiolitis was defined as the first episode of wheezing with respiratory tract symptoms in children younger than 12 months or the second episode without a personal or family history of asthma or atopy, according to national and international guidelines.^18,19^ We excluded infants with previous immunization with palivizumab or maternal vaccination against RSV during pregnancy, those with a personal history of two or more wheezing episodes (considered as asthma), those who received nirsevimab before the start of the immunization campaign. We also excluded infants with incomplete data on nirsevimab immunization status or results of RSV test.

Case patients were defined as children who had been hospitalized for bronchiolitis with a positive RSV test. Control patients were defined as children who had been hospitalized for bronchiolitis with a negative RSV test.

### 1.2. RSV immunization campaign in France

In winter 2023-2024 period, France was one of the first countries to implement a national immunization campaign using nirsevimab for all infants during their first epidemic season of RSV. For the first RSV season following the implementation of nirsevimab, French authorities issued a recommendation for a single dose of nirsevimab between September 15, 2023 and the end of the RSV season, for all infants born after February 6, 2023. For the second RSV season, the French guidelines recommended two strategies, which were implemented on September 15, 2024. First, a single dose of nirsevimab can be administrated for all infants born after January 1, 2024. Secondly, the maternal vaccine against RSV using RSVpreF can be administered between 32 and 36 weeks of gestation to increase the passive immunization in infants.

### 1.3. RSV testing and genome sequencing

A nasopharyngeal swab was obtained from each patient for an RSV diagnostic test, consisting of a polymerase chain reaction (PCR) assay or a rapid antigen test. Among the RSV-positive samples, we selected with 1:1 ratio nirsevimab-exposed infants with post-exposure RSV infection and non-exposed infants with RSV infection for whole genome sequencing.^13^ Providing a similar proportion of non-immunized and immunized children among selected cases to be whole genome sequenced to optimize statistical power to describe the RSV genomic diversity in each group. This selection was performed blinded of the clinical characteristics of children except nirsevimab status.

We analyzed consensus sequences together with a contextual reference dataset selected from Goya et al to ensure good representativeness of RSV diversity. We computed a multiple sequence alignment from these sequences using MAFFT (version 7.505) and phylogenies were inferred using IQ-TREE (version 2.2.5; maximum likelihood; general time reversible model, 1000 bootstrap samples). The distribution of lineages over the first two seasons of the immunization campaign was determined using these sequenced RSV genome samples. RSV lineages were assigned according to the standardized RGCC nomenclature proposed by Goya et al.^20^ Consensus lineage-defining amino acid signatures of A.D.3 lineage and its descendants were reported from the corresponding lineage designation table and are summarized in Table S1, Supplementary Appendix.

RSV lineages were grouped into six groups according to their genotypic proximity as follows: A.D.1, A.D.1.5 and A.D.1.6; A.D.3, A.D.3.1 and A.D.3.3; A.D.4; A.D.5, A.D.5.1 and A.D.5.2; B.D.4.1 and B.D.4.1.1; B.D.E.1, B.D.E.1.1 and B.D.E.1.2.^20^

RSV strains from a subset of RSV-positive samples from the POLYRES study that could be successfully cultured on HEp-2 cells were isolated and amplified as previously described. The susceptibility of these isolates to nirsevimab was assessed by a conventional plaque reduction neutralisation assay. Briefly, plaque-forming units of RSV were incubated with serial dilutions of nirsevimab before titration. Plaque counts were plotted against nirsevimab concentrations to estimate the half-maximal inhibitory concentration (IC50). The RSV Long strain was included as a reference virus in each experiment as previously described. Fold changes (FCs) in IC50 were calculated for each isolate relative to the RSV Long reference strain.^13^

### 1.4. Setting and data collection

This study was based on data from three national multicenter prospective studies which were named OVNI^9^, ENVIE^8^ and POLYRES.^13^ We collected data on demographic characteristics, comorbidities at inclusion (bronchopulmonary dysplasia, congenital heart disease, immune deficiencies, neurological conditions), RSV test results, nirsevimab immunization status and severity of bronchiolitis (hospitalization and ventilatory support). The 12 participating centers were secondary or tertiary hospital with a pediatric department located in France.

### 1.5. Outcome and exposure measures

The primary outcome was RSV-positive bronchiolitis in infants younger than 12 months according to lineages. The exposure was nirsevimab immunization status among both case and control patients.

### 1.6. Sample size calculation

On the basis of an expected nirsevimab coverage of 50% in the RSV-negative population of infants younger than 12 months in France, the study sample size was calculated to detect a 50% reduction in the odds of nirsevimab immunization among case patients versus control patients according to RSV lineages. Assuming a two-sided α of 0.05 and a power of 0.80, we needed to include 95 patients in each of the five predominant lineage group plus 95 control patients (i.e., 475 case patients and 95 control patients in total).

### 1.7. Statistical analyses

Descriptive results were presented as median (interquartile range, Q1-Q3) for continuous variables, and as total numbers (percentages) for categorical variables.

Due to the random selection process of RSV strains to be whole genome sequenced based on nirsevimab status and taking into account the major influence of RSV season on circulating lineages, we stratified the multiple imputation of missing data by these two parameters. Thus, we performed four separate multiple imputations by chained equations (first season and immunized by nirsevimab, first season and non-immunized by nirsevimab, second season and immunized by nirsevimab, second season and non-immunized by nirsevimab) using m= 20 imputations and twenty iterations. The RSV lineage variable was imputed using random forest method with six predictors as age, sex, gestational age at birth (weeks), birth weight (g), comorbidities and month of diagnosis. Moreover, we performed multiple imputation by chained equations with five iterations using predictive mean matching to handle missing data in all other binary or continuous variables included in the multivariable analyses. The results were combined using Rubin’s rules.^21,22^

The effectiveness of nirsevimab against hospitalization for RSV bronchiolitis was estimated for each RSV lineage group using multivariable mixed-effects logistic regression, comparing cases to controls according to nirsevimab immunization status. Models were adjusted for age, sex, birth weight, gestational age at birth, comorbidities as defined above, RSV season, and month of diagnosis (grouped by 2 months), considered as potential confounders and included a random effect for centers. The effectiveness was calculated as: effectiveness = 100% × (1 − adjusted odds ratio (OR)).

The risk of nirsevimab immunization failure across RSV lineages was assessed by comparing adjusted odds ratios to a reference lineage. The reference lineage group, B.D.4.1 and B.D.4.1.1 group, was defined because it was the lineage with the highest effectiveness. The RSV lineage A.D.4 was excluded from lineages analyses due to a total number of fewer than ten patients. Sensitivity analyses were performed to assess the robustness of the study findings. First, we assessed the effectiveness of nirsevimab against hospitalization for RSV-bronchiolitis using the Heckman selection model to impute missing lineage data and account for potential selection bias. In these models, RSV sequencing was modelled in a probit selection equation and RSV lineage in an outcome equation, with correlated error terms and inclusion of the inverse Mills ratio to allow imputation under a missing not at random mechanism.^23^ We included age, sex, PICU admission, RSV season and nirsevimab immunization status variables in the Heckman’s model. Second, we used the same Heckman selection model to estimate the risk of nirsevimab immunization failure for each lineage by comparing adjusted odds ratios to a reference lineage as previously described in the main analysis. Third, we performed a multivariable analysis to estimate the risk of nirsevimab immunization failure for each lineage without imputation of missing lineage data. All patients who had an RSV-positive test without RSV sequencing were excluded from this analysis. Fourth, we performed a multivariable analysis without considering the 7-day delay between nirsevimab immunization and hospitalization. Fifth, we conducted a multivariable analysis stratified by epidemic RSV season. Sixth, we performed an unadjusted analysis without a random effect for centers using a logistic regression.

All the statistical tests were two-sided, with a significance level of p < 0.05. Analyses were performed with R software version 4.5.2.

### 1.8. Ethical approval

The study was approved by the French National Commission on Informatics and Liberty (no. 1921226) and by an ethics committee (CHI Creteil Hospital, France for OVNI and ENVIE; CH Henri Mondor, France for POLYRES).

## 2. RESULTS

### 2.1. Population characteristics

During the study period, 3365 infants were hospitalized for bronchiolitis. Among them, 346 (11.5%) were excluded (Figure 1) and their general characteristics are detailed in Table S2, Supplementary Appendix. We included 3019 patients with bronchiolitis in the final analysis with 2273 case patients and 746 control patients. Clinical characteristics are detailed in Table 1. Median age was 3.2 months (Q1-Q3, 1.6–5.5) among the case patients and 2.8 months (Q1-Q3, 1.3–5.6) among the control patients. The general characteristics were similar between patients with and without RSV whole-genome sequencing, except for nirsevimab immunization status, given the sampling approach used for sequencing as described in Methods section (Table S3, Supplementary Appendix). Moreover, the distribution of RSV lineages before and after imputation using multiple imputation chained equations were similar (Table S4, Supplementary Appendix).

**Table 1.**
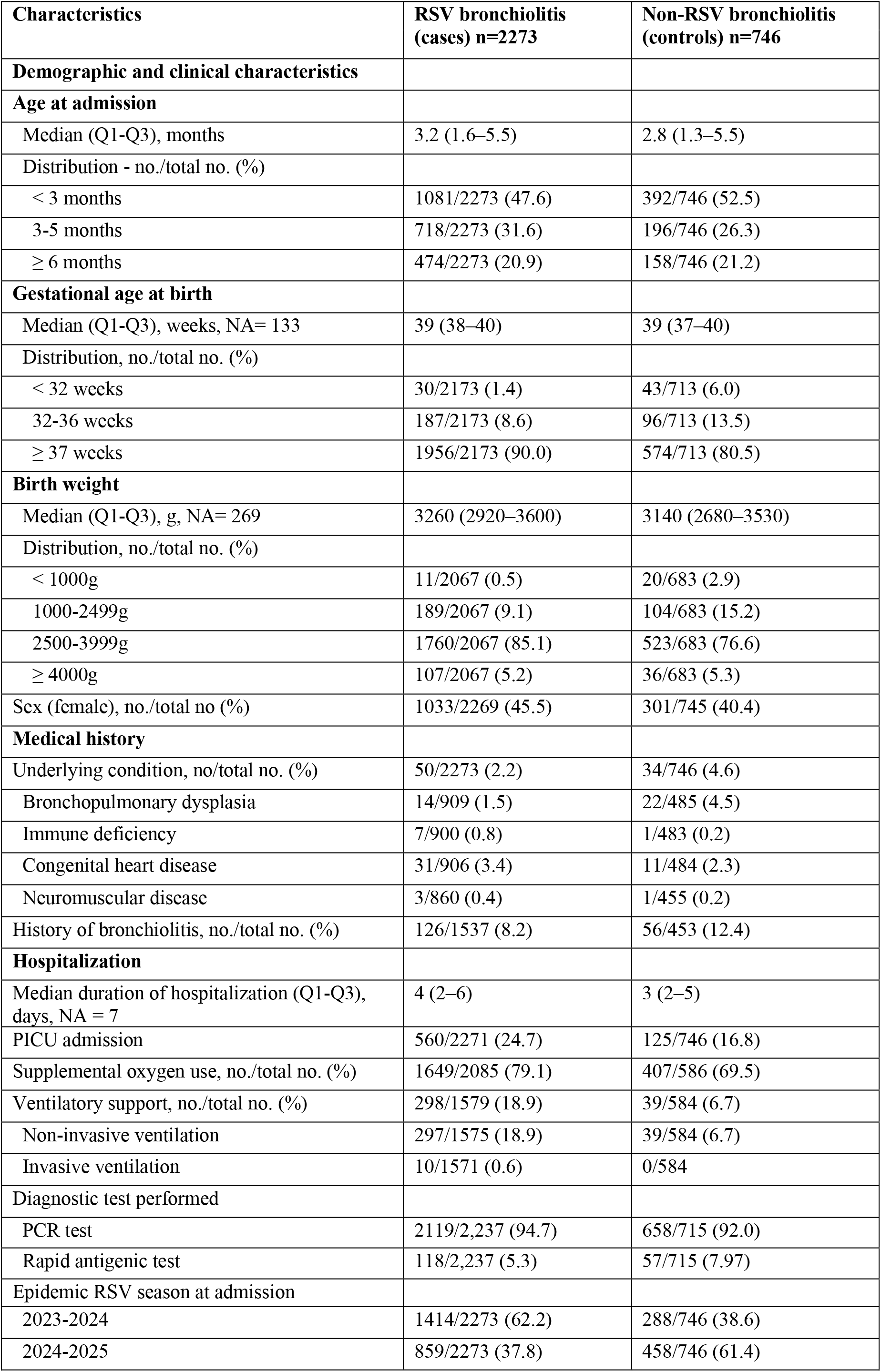
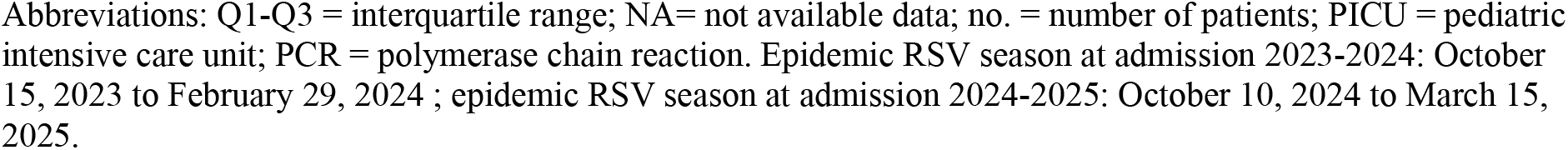
Patient characteristics of the study population (n= 3019).

| Characteristics | RSV bronchiolitis<br>(cases) n=2273 | Non-RSV bronchiolitis<br>(controls) n=746 |
| --- | --- | --- |
| **Demographic and clinical characteristics** |  |  |
| **Age at admission** |  |  |
| Median (Q1-Q3), months | 3.2 (1.6–5.5) | 2.8 (1.3–5.5) |
| Distribution - no./total no. (%) |  |  |
| < 3 months | 1081/2273 (47.6) | 392/746 (52.5) |
| 3-5 months | 718/2273 (31.6) | 196/746 (26.3) |
| ≥ 6 months | 474/2273 (20.9) | 158/746 (21.2) |
| **Gestational age at birth** |  |  |
| Median (Q1-Q3), weeks, NA= 133 | 39 (38–40) | 39 (37–40) |
| Distribution, no./total no. (%) |  |  |
| < 32 weeks | 30/2173 (1.4) | 43/713 (6.0) |
| 32-36 weeks | 187/2173 (8.6) | 96/713 (13.5) |
| ≥ 37 weeks | 1956/2173 (90.0) | 574/713 (80.5) |
| **Birth weight** |  |  |
| Median (Q1-Q3), g, NA= 269 | 3260 (2920–3600) | 3140 (2680–3530) |
| Distribution, no./total no. (%) |  |  |
| < 1000g | 11/2067 (0.5) | 20/683 (2.9) |
| 1000-2499g | 189/2067 (9.1) | 104/683 (15.2) |
| 2500-3999g | 1760/2067 (85.1) | 523/683 (76.6) |
| ≥ 4000g | 107/2067 (5.2) | 36/683 (5.3) |
| Sex (female), no./total no (%) | 1033/2269 (45.5) | 301/745 (40.4) |
| **Medical history** |  |  |
| Underlying condition, no/total no. (%) | 50/2273 (2.2) | 34/746 (4.6) |
| Bronchopulmonary dysplasia | 14/909 (1.5) | 22/485 (4.5) |
| Immune deficiency | 7/900 (0.8) | 1/483 (0.2) |
| Congenital heart disease | 31/906 (3.4) | 11/484 (2.3) |
| Neuromuscular disease | 3/860 (0.4) | 1/455 (0.2) |
| History of bronchiolitis, no./total no. (%) | 126/1537 (8.2) | 56/453 (12.4) |
| **Hospitalization** |  |  |
| Median duration of hospitalization (Q1-Q3), days, NA = 7 | 4 (2–6) | 3 (2–5) |
| PICU admission | 560/2271 (24.7) | 125/746 (16.8) |
| Supplemental oxygen use, no./total no. (%) | 1649/2085 (79.1) | 407/586 (69.5) |
| Ventilatory support, no./total no. (%) | 298/1579 (18.9) | 39/584 (6.7) |
| Non-invasive ventilation | 297/1575 (18.9) | 39/584 (6.7) |
| Invasive ventilation | 10/1571 (0.6) | 0/584 |
| Diagnostic test performed |  |  |
| PCR test | 2119/2,237 (94.7) | 658/715 (92.0) |
| Rapid antigenic test | 118/2,237 (5.3) | 57/715 (7.97) |
| Epidemic RSV season at admission |  |  |
| 2023-2024 | 1414/2273 (62.2) | 288/746 (38.6) |
| 2024-2025 | 859/2273 (37.8) | 458/746 (61.4) |

Abbreviations: Q1-Q3 = interquartile range; NA= not available data; no. = number of patients; PICU = pediatric intensive care unit; PCR = polymerase chain reaction. Epidemic RSV season at admission 2023-2024: October 15, 2023 to February 29, 2024 ; epidemic RSV season at admission 2024-2025: October 10, 2024 to March 15, 2025.

**Figure 1:**
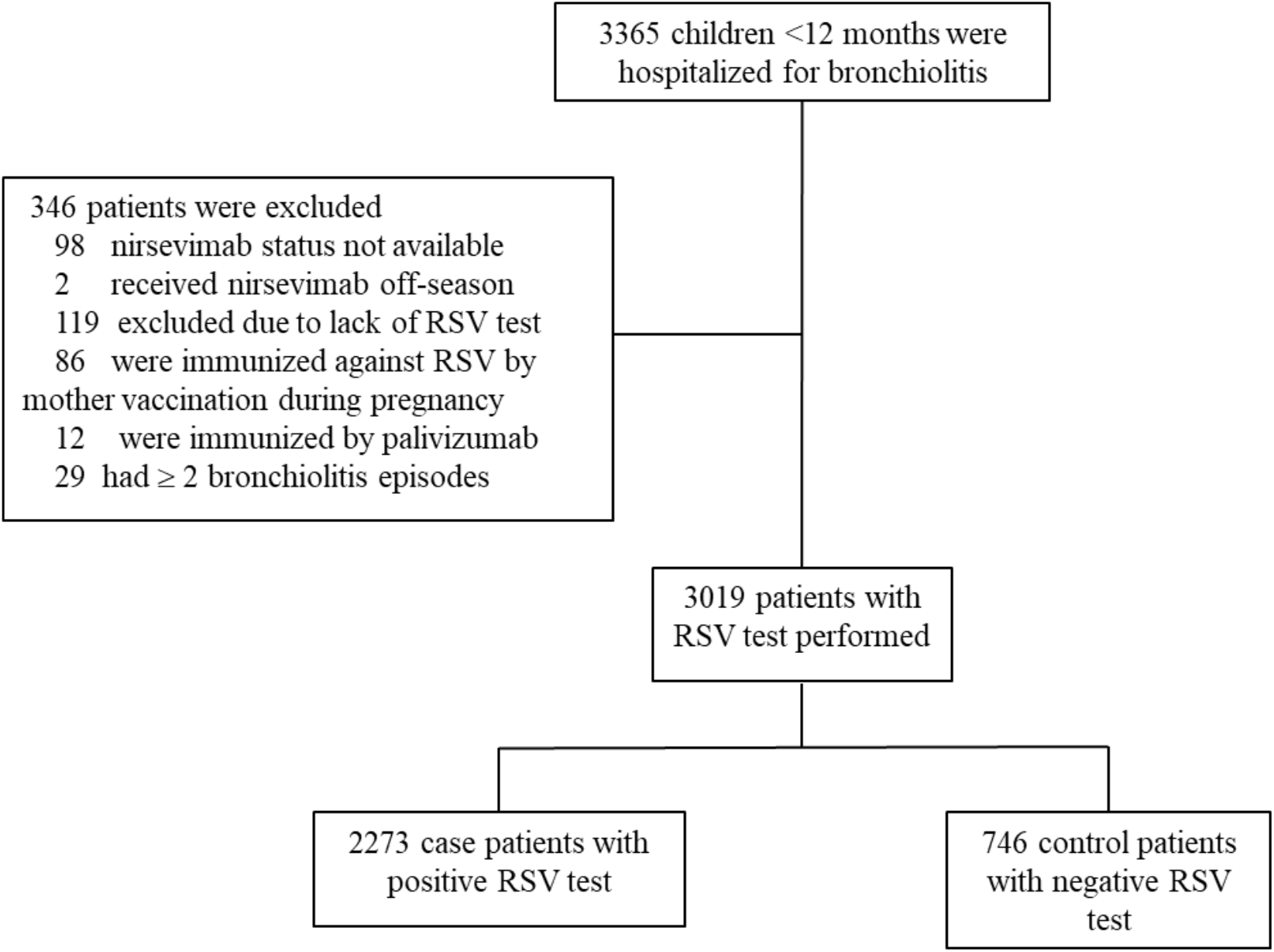
flowchart of the study. Abbreviations: RSV= respiratory syncytial virus.

### 2.2. Outcomes

In the study population, 373/2273 (16.4%) case patients received nirsevimab versus 423/746 (56.7%) control patients. The distribution of RSV lineages before and after imputation using multiple imputation chained equations is presented in Table 2. The overall adjusted effectiveness of nirsevimab against hospitalization for RSV-bronchiolitis was 82.1% (95% CI, 77.8–85.6). The adjusted effectiveness of nirsevimab by RSV subtype was 79.4% (95% CI, 73.7–83.8) against RSV-A and 88.4% (95% CI, 84.5–91.4) against RSV-B. According to RSV lineages, the highest effectiveness of nirsevimab was 90.0% (95% CI, 83.8–93.8) against B.D.4.1 lineages and the lowest effectiveness of nirsevimab was 67.8% (95% CI, 56.0–76.5) against A.D.3 lineages (Figure 2). Compared with B.D.4.1 lineages, A.D.3 lineages were associated with a 3.2-fold higher odds of immunization failure (OR, 3.2; 95% CI 1.8–5.7) (Figure 3). Among children with breakthrough infection, the interval between immunization and hospitalization were similar across RSV lineages. Rates of respiratory viral co-infections were similar between A.D.3 lineages and other lineages (Tables S5 & S6, Supplementary Appendix). All sensitivity analyses showed similar results (Figure S1 to S5, Supplementary Appendix). RSV isolates susceptibility testing to nirsevimab according to lineages are presented in Figure S8, Supplementary Appendix. The highest median IC50 fold-change relative to the reference strain was observed for the A.D.3 lineage (1.48; range, 0.32–4.28). None of the tested isolates harboured known nirsevimab resistance-associated substitutions.

**Table 2.** Distribution of RSV lineages using MICE imputation model – no. (%)

| RSV season | RSV lineages | Before MICE | After MICE |
| --- | --- | --- | --- |
| 2023-2024 | A.D.1, A.D.1.5 and A.D.1.6 | 52 (46) | 648 (46) |
|  | A.D.3, A.D.3.1 and A.D.3.3 | 14 (12) | 88 (6) |
|  | A.D.4 | 1 (1) | 1 (0) |
|  | A.D.5, A.D.5.1 and A.D.5.2 | 27 (24) | 355 (25) |
|  | B.D.4.1 and B.D.4.1.1 | 7 (6) | 105 (7) |
|  | B.D.E.1, B.D.E.1.1 and B.D.E.1.2 | 11 (10) | 217 (15) |
| 2024-2025 | A.D.1, A.D.1.5 and A.D.1.6 | 22 (19) | 172 (20) |
|  | A.D.3, A.D.3.1 and A.D.3.3 | 36 (31) | 249 (29) |
|  | A.D.4 | 0 (0) | 0 (0) |
|  | A.D.5, A.D.5.1 and A.D.5.2 | 3 (3) | 28 (3) |
|  | B.D.4.1 and B.D.4.1.1 | 13 (11) | 91 (11) |
|  | B.D.E.1, B.D.E.1.1 and B.D.E.1.2 | 44 (37) | 319 (37) |

Abbreviations: RSV = respiratory syncytial virus. RSV season 2023-2024: October 15, 2023 to February 29, 2024; RSV season 2024-2025: October 10, 2024 to March 15, 2025.

**Figure 2.**
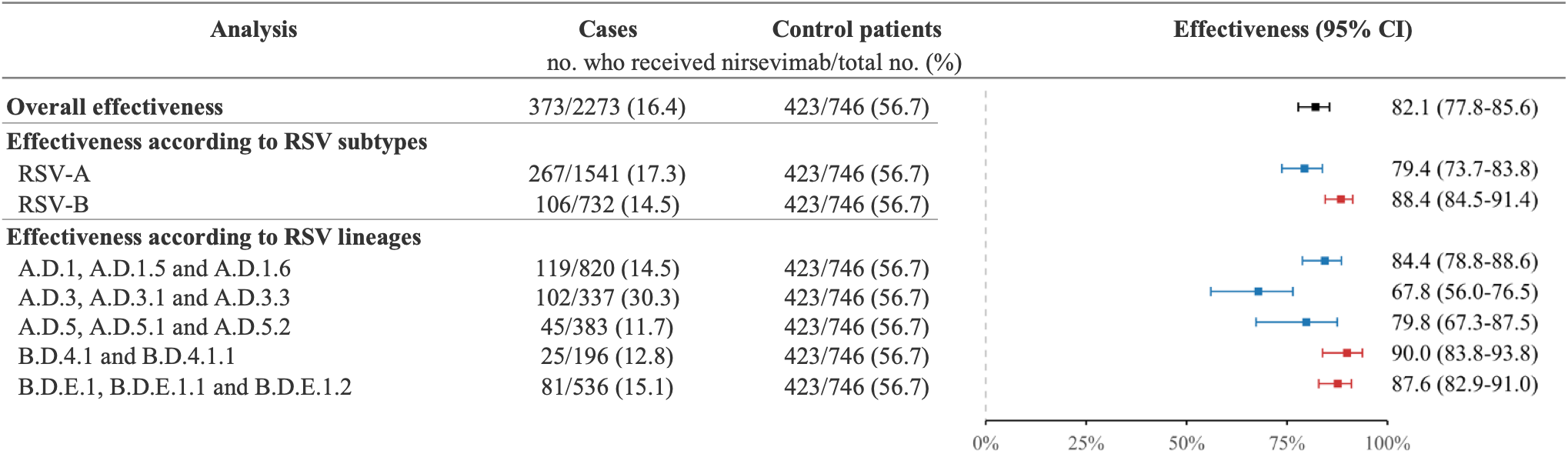
Effectiveness of nirsevimab against hospitalization for RSV bronchiolitis in infants according to RSV groups and lineages (n= 3019). Case patients were those with RSV-positive bronchiolitis younger than 12 months. Control patients were those with RSV-negative bronchiolitis. Effectiveness was calculated as 100% × (1 − adjusted odds ratio). The multivariable logistic regression model was adjusted for age, sex, birth weight, gestational age at birth, comorbidities, RSV season, and month of diagnosis (grouped by 2 months) and included a random effect for centers.

**Figure 3.**
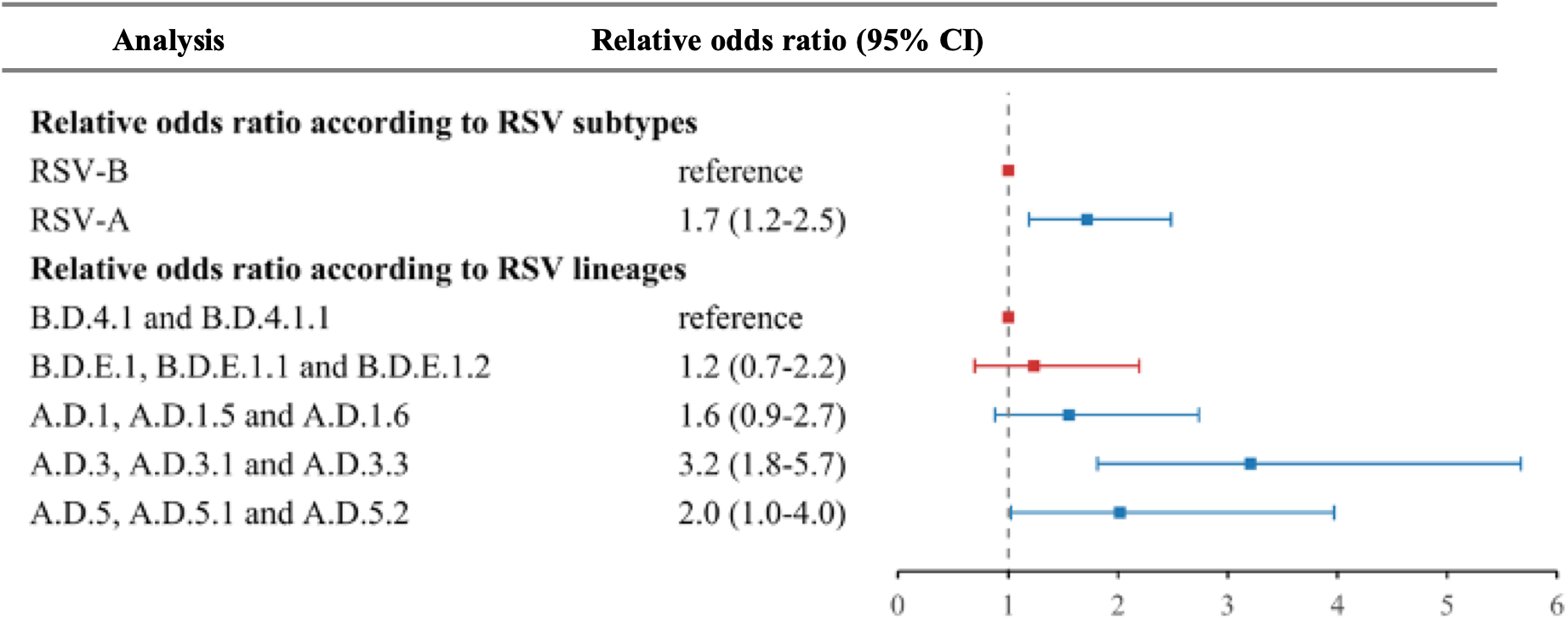
Risk of nirsevimab immunization failure according to RSV groups and lineages (n= 3017). Risk of nirsevimab immunization failure was estimated by comparing adjusted odds ratios to a reference lineage. The multivariable logistic regression model was adjusted for age, sex, birth weight, gestational age at birth, comorbidities, RSV season, month of diagnosis (grouped by 2 months) and included a random effect for centers.

## 3. DISCUSSION

To the best of our knowledge, this real-world study is the first to assess the effectiveness of nirsevimab against hospitalized RSV bronchiolitis according to RSV subtypes and lineages. This study showed a higher effectiveness of nirsevimab against hospitalization for bronchiolitis with RSV-B than with RSV-A, respectively 88.4% (95% CI, 84.5–91.4) and 79.4% (95% CI, 73.7–83.8). We also found a significantly higher risk of immunization failure with lineages derived from A.D.3 than with those derived from B.D.4.1 (OR, 3.2; 95% CI 1.8–5.7).

Changes in the distribution of lineages were observed in France across the two seasons, with an increase in the circulation of lineages derived from RSV-B in the 2024-2025 RSV season.^24^ In addition, a higher degree of genetic variability was observed in RSV-B strains than in RSV-A strains, particularly with regard to substitutions identified in the binding site of nirsevimab.^25^ Recently, *Fourati et al*. described rare cases of RSV-B breakthrough infections with resistance-associated substitutions.^13,14^ Despite these substitutions described in RSV-B, the effectiveness of nirsevimab against RSV remains high during the circulation of RSV-B in the 2024-2025 RSV season.^16,17^ Our findings were consistent with a high effectiveness of nirsevimab against RSV-B strains and mitigate concerns about rapid expansion of nirsevimab resistant RSV strains. In contrast, we observed a lower effectiveness of nirsevimab against hospitalization for RSV bronchiolitis derived from the RSV-A A.D.3 lineage and its sublineages (including A.D.3, AD.3.1 and A.D.3.3). In our study, the interval between nirsevimab immunization and hospitalization did not differ across RSV lineages suggesting that the lower effectiveness observed for A.D.3 lineages was not due to wanning immunity. Moreover, viral co-infections were not more frequent with A.D.3 lineages versus other lineages, suggesting that co-infections with other viruses are unliked to explain our results. Consistent with our clinical observations, preliminary phenotypic analyses from POLYRES study suggest higher IC_50_ values for A.D.3 lineages relative to other RSV lineages although these differences were not statistically significant (Figure S8, Supplementary Appendix).^14^ Of note, genotypic studies have not evidenced any resistance-associated substitutions to nirsevimab within the A.D.3 lineage.^13,25^ Furthermore, a recent study analyzed phenotypic divergence of RSV isolates and reported lower neutralization of several RSV-A isolates compared with RSV-B isolates despite the absence of substitutions affecting the known nirsevimab binding site.^26^ These observations raises the possibility that mechanisms other than direct modifications of the nirsevimab epitope may contribute to reduced susceptibility. As a distinct RSV lineage, RSV-A A.D.3 is defined by a unique combination of lineage-defining amino acid substitutions distributed across multiple viral proteins, including the F and G glycoproteins (Supplementary Table S1, Supplementary Appendix). We speculate that such lineage-specific changes could modify the structural organization the prefusion F protein, and, consequently, the accessibility of antigenic site Ø on the protein.^27^ Further phenotypic studies using physiologically relevant three-dimensional human airway epithelial models are needed to determine whether the A.D.3 genetic background directly affects susceptibility to nirsevimab. In parallel, larger real-world effectiveness studies conducted in independent populations will be essential to confirm our findings. Importantly, during the peak of 2025-2026 epidemic RSV season, A.D.3 lineages were predominant among RSV strains circulating in France with around 65% of RSV samples.^24^ During the same period, a decrease of effectiveness of nirsevimab against RSV bronchiolitis in primary care settings was reported,^28^ which is consistent with our findings of a reduced effectiveness for this specific lineage. Further studies from other settings are required to confirm population level impact of these findings.

Whether the widespread implementation of nirsevimab in infants since 2023 has driven the selection of the A.D.3 lineage remains uncertain. RSV also circulate in older children and adults, who are not targeted by nirsevimab immunization campaign but still contribute to community transmission.^29,30^ Furthermore, the proportion of RSV strains from the A.D.3 lineages has increased in Africa and Southeast Asia since 2025, despite these regions not having been exposed to nirsevimab.^24^ Consequently, the selective pressure imposed by nirsevimab on circulating RSV strains may be limited, though it should be monitored over coming years.

Our study has several limitations. First, due to the observational case-control study design, uncontrolled confounders cannot be excluded and causative conclusions cannot be drawn. However, the test-negative design is widely used to assess the vaccine effectiveness and more recently to assess the real-world effectiveness of nirsevimab^16,17,31^. Because case and control patients were both admitted for bronchiolitis, the risk of bias related to health-seeking behaviors may be limited. Second, due to the POLYRES study design which aimed to select one breakthrough infection for each non-immunized RSV case selected for whole-genome sequencing, the proportion of immunized children among sequenced RSV cases was overrepresented compared with the source population. To address this, we performed multiple imputation by chained equations stratified by the nirsevimab immunization status, preserving the immunization coverage of the overall cohort and reducing the potential for selection bias. Third, we performed a multiple imputation by chained equation which are usually applied to variables with a low proportion of missing data. Moreover, these imputation methods are formally validated under the missing-at-random assumption. However, the decision to send RSV samples for whole-genome sequencing was only based on the nirsevimab immunization status, as detailed in the Methods section, and was blinded to the children’s clinical characteristics of the children, a missing non at random mechanism is unlikely. Furthermore, the results after imputation using Heckman’s selection model and without imputation showed similar results, reinforcing the robustness of our findings. Fourth, because some patients did not have a PCR test and RSV detection was based on rapid antigen testing, we cannot exclude false-negative or false-positive cases^32^.

## 4. CONCLUSION

This real-world study evaluated the effectiveness of nirsevimab against hospitalization for RSV-bronchiolitis according to RSV subtypes and lineages during the two initial RSV seasons following the nirsevimab implementation in France. Despite the widespread use of nirsevimab, the overall effectiveness remained high. We observed differences in effectiveness across RSV lineages, particularly a decrease in nirsevimab effectiveness against lineages derived from the A.D.3 lineage although the underlying mechanisms remain unclear. These results highlight the importance of active molecular and clinical surveillance in the coming years as nirsevimab use expands.

## Data Availability

All data produced in the present work are contained in the manuscript

