## Supplementary material for "Effectiveness of nirsevimab against hospitalization for RSV bronchiolitis according to RSV lineages"

**Table S1. Defining substitutions of RSV lineages.**

**Table S2. Characteristics of included and excluded patients.**

**Table S3. Characteristics of patients hospitalized for RSV-bronchiolitis with or without RSV genome sequencing.**

**Table S4. Distribution of RSV lineages using Heckman's imputation model.**

**Table S5. Viral co-infections according to RSV lineages in the study population.**

**Table S6. Time between immunization with nirsevimab and hospitalization according to RSV lineages in the study population.**

**Figure S1. Effectiveness of nirsevimab against hospitalization for RSV bronchiolitis in infants using Heckman's model.**

**Figure S2. Risk of nirsevimab immunization failure according to RSV groups and lineages using Heckman's model.**

**Figure S3. Risk of nirsevimab immunization failure according to RSV groups and lineages without imputation (n=230).**

**Figure S4. Effectiveness of nirsevimab against hospitalization for RSV bronchiolitis in infants, excluding those were immunised less than 7 days before hospitalization.**

**Figure S5. Effectiveness of nirsevimab against hospitalization for RSV bronchiolitis in infants in 2023-2024 epidemic RSV season.**

**Figure S6. Effectiveness of nirsevimab against hospitalization for RSV bronchiolitis in infants in 2024-2025 epidemic RSV season.**

**Figure S7. Unadjusted effectiveness of nirsevimab against hospitalization for RSV bronchiolitis in infants (n=3019).**

**Figure S8. Nirsevimab susceptibility of RSV isolates.**

**Table S1. Defining substitutions of RSV A.D.3, A.D.3.1 and A.D.3.3 sublineages.**

| | **defining substitutions (compared to parental A.D)** |
|---|---|
| A.D.3 | F:12I, G:113I, L:835M |
| A.D.3.1 | F:12I, F:20F, F:276N, G:101S, G:113I, L:835M |
| A.D.3.3 | F:12I, G:113I, G:204N, L:835M, M2-2:45Y, NS1:2D |

**Table S2. Characteristics of included and excluded patients.**

| **Characteristics** | **Included patients (n= 3019)** | **Excluded patients (n= 346)** |
|---|---|---|
| **Demographic and clinical characteristics** | | |
| Age at admission - median (Q1-Q3), months. | 3.1 (1.6–5.5) | 3.5 (1.3–6.5) |
| Gestationnal age at birth - median (Q1-Q3), GA | 39 (38–40) | 39 (38–40) |
| NA= 158 | | |
| Birth weight - median (Q1-Q3), g | 3232.5 (2860–3585) | 3240 (2860–3555) |
| NA= 318 | | |
| Sex (female), no./total no (%) | 1334/3014 (44.3) | 149/346 (43.1) |
| Positive RSV test, no./total no (%) | 2,273/3,019 (75.3%) | 120/204 (58.8%) |
| Nirsevimab immunization | 796/3,019 (26.4%) | 55/248 (22.2%) |
| **Medical history** | | |
| Underlying condition, no/total no. (%) | 84/3019 (2.8) | 18/346 (5.2) |
| History of bronchiolitis, no./total no. (%) | 182/1990 (9.2) | 58/254 (22.8) |
| **Hospitalization** | | |
| Median duration of hospitalization (Q1-Q3), days, NA = | 4 (2–6) | 3 (1–5) |
| PICU admission | 685/3017 (22.7) | 57/346 (16.5) |
| Supplemental oxygen use, no./total no. (%) | 2056/2671 (77.0) | 202/296 (68.2) |
| Ventilatory support, no./total no. (%) | 337/2163 (15.6) | 24/293 (8.2) |
| Epidemic season at admission | | |
| 2023-2024 | 1702/3019 (56.4) | 89/346 (25.7) |
| 2024-2025 | 1317/3019 (43.6) | 257/346 (74.3) |

**Table S3. Characteristics of patients hospitalized for RSV-bronchiolitis with or without RSV genome sequencing.**

| Characteristics | Unsequenced RSV samples (n= 2043) | Sequenced RSV samples (n= 230) |
|---|---|---|
| **Demographic and clinical characteristics** | | |
| Age at admission - median (Q1-Q3), months. | 3.3 (1.7–5.6) | 2.3 (1.2–4.1) |
| Gestational age at birth - median (Q1-Q3), GA | 39 (38–40) | 39 (38–40) |
| NA= 100 | | |
| Birth weight - median (Q1-Q3), g | 3270 (2910–3600) | 3210 (2940–3610) |
| NA= 206 | | |
| Sex (female), no./total no (%) | 918/2039 (45.0) | 115/230 (50.0) |
| Nirsevimab immunization | 278/2043 (13.6) | 95/230 (41.3) |
| **Medical history** | | |
| Underlying condition, no/total no. (%) | 45/2043 (2.2) | 5/230 (2.2) |
| History of bronchiolitis, no./total no. (%) | 117/1377 (8.5) | 9/160 (5.6) |
| **Hospitalization** | | |
| Median duration of hospitalization (Q1-Q3), | 4 (2–6) | 4 (3–6) |
| PICU admission | 504/2041 (24.7) | 56/230 (24.3) |
| Supplemental oxygen use, no./total no. (%) | 1480/1873 (79.0) | 169/212 (79.7) |
| Ventilatory support, no./total no. (%) | 275/1401 (19.6) | 23/178 (12.9) |
| Epidemic season at admission | | |
| 2023-2024 | 1302/2043 (63.7) | 112/230 (48.7) |
| 2024-2025 | 741/2043 (36.3) | 118/230 (51.3) |

**Table S4. Distribution of RSV lineages using Heckman's imputation model – no. (%)**

| RSV season | RSV lineages | Before imputation | After imputation |
|---|---|---|---|
| 2023-2024 | A.D.1, A.D.1.5 and A.D.1.6 | 52 (46.4%) | 681.8 (48.3%) |
| | A.D.3, A.D.3.1 and A.D.3.3 | 14 (12.5%) | 128.2 (9.1%) |
| | A.D.4 | 1 (0.9%) | - |
| | A.D.5, A.D.5.1 and A.D.5.2 | 27 (24.1%) | 368.8 (26.1%) |
| | B.D.4.1 and B.D.4.1.1 | 7 (6.2%) | 78.6 (5.6%) |
| | B.D.E.1, B.D.E.1.1 and B.D.E.1.2 | 11 (9.8%) | 155.6 (11.0%) |
| 2024-2025 | A.D.1, A.D.1.5 and A.D.1.6 | 22 (18.6%) | 162.6 (18.9%) |
| | A.D.3, A.D.3.1 and A.D.3.3 | 36 (30.5%) | 247.4 (28.8%) |
| | A.D.4 | 0 (0%) | - |
| | A.D.5, A.D.5.1 and A.D.5.2 | 3 (2.5%) | 32.0 (3.7%) |
| | B.D.4.1 and B.D.4.1.1 | 13 (11.0%) | 90.0 (10.5%) |
| | B.D.E.1, B.D.E.1.1 and B.D.E.1.2 | 44 (37.3%) | 327.0 (38.1%) |

**Table S5. Viral co-infections according to RSV lineages in the study population – no. (%).**

| RSV lineages | ≥ 1 virus identified other than RSV |
|---|---|
| A.D.1, A.D.1.5 and A.D.1.6 | 20/72 (28) |
| A.D.3, A.D.3.1 and A.D.3.3 | 12/46 (26) |
| A.D.5, A.D.5.1 and A.D.5.2 | 4/29 (14) |
| B.D.4.1 and B.D.4.1.1 | 5/20 (25) |
| B.D.E.1, B.D.E.1.1 and B.D.E.1.2 | 12/50 (24) |

**Table S6. Time between immunization with nirsevimab and hospitalization according to RSV lineages in the study population – days (Q1-Q3) (n= 2271).**

| RSV lineages | Overall | 2023-2024 RSV season | 2024-2025 RSV season |
|---|---|---|---|
| A.D.1, A.D.1.5 and A.D.1.6 | 37.0 (23.0–49.0) | 35.0 (23.0–44.5) | 66.0 (26.5–79.0) |
| A.D.3, A.D.3.1 and A.D.3.3 | 43.0 (28.5–66.0) | 36.5 (18.8–54.5) | 46.0 (33.0–73.0) |
| A.D.5, A.D.5.1 and A.D.5.2 | 35.0 (21.0–47.0) | 35.0 (21.0–47.0) | - |
| B.D.4.1 and B.D.4.1.1 | 54.5 (38.0–75.8) | 9.0 (7.0–23.0) | 62.0 (44.5–81.0) |
| B.D.E.1, B.D.E.1.1 and B.D.E.1.2 | 42.0 (18.0–63.5) | 29.0 (26.5–42.0) | 42.0 (18.0–65.0) |

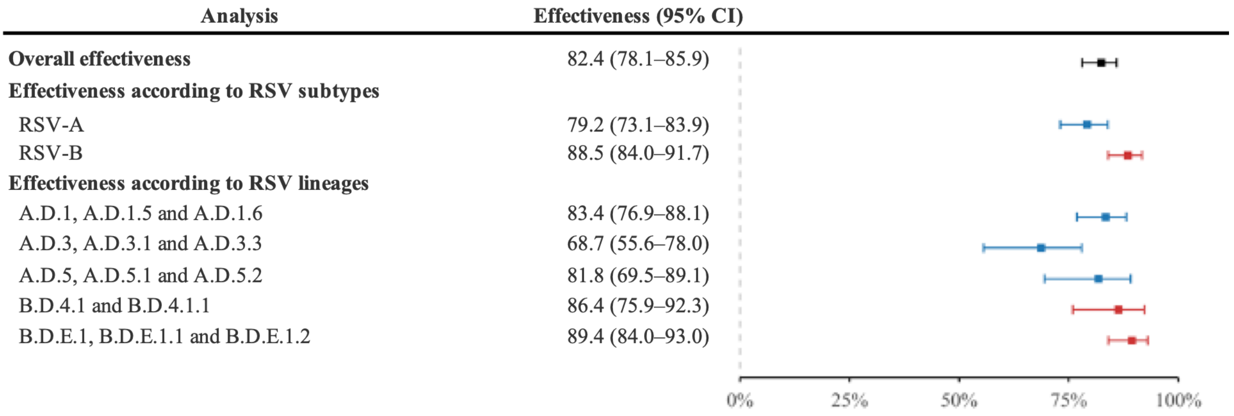


**Figure S1. Effectiveness of nirsevimab against hospitalization for RSV bronchiolitis in infants using Heckman's model.**

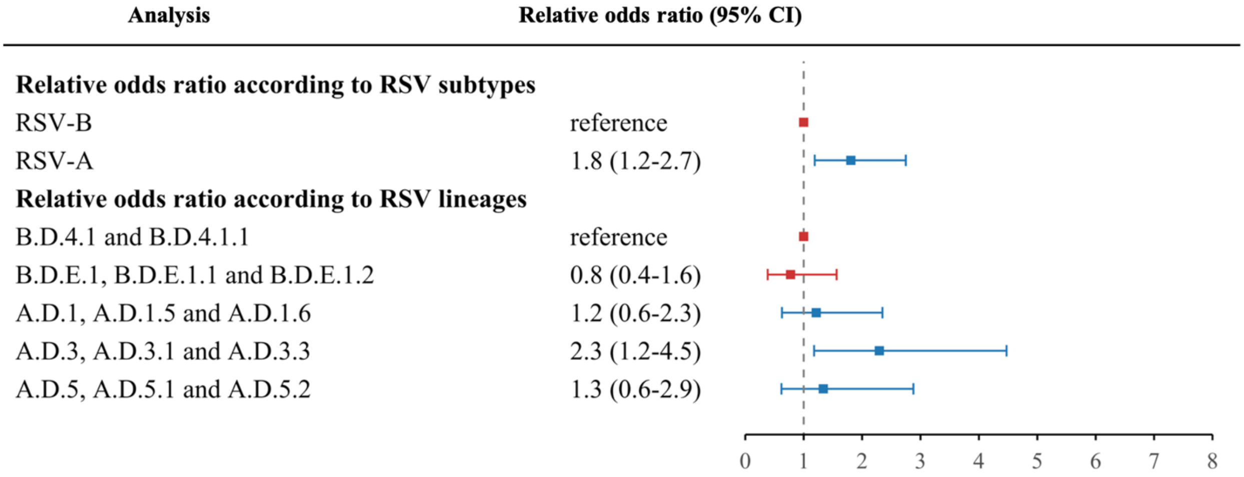


**Figure S2. Risk of nirsevimab immunization failure according to RSV groups and lineages using Heckman's model.**

| Analysis | Relative odds ratio (95% CI) |
|---|---|
| **Relative odds ratio according to RSV subtypes** | |
| RSV-B | reference |
| RSV-A | 2.5 (1.2-5.2) |
| **Relative odds ratio according to RSV lineages** | |
| B.D.4.1 and B.D.4.1.1 | reference |
| B.D.E.1, B.D.E.1.1 and B.D.E.1.2 | 0.7 (0.2-2.2) |
| A.D.1, A.D.1.5 and A.D.1.6 | 1.7 (0.5-5.5) |
| A.D.3, A.D.3.1 and A.D.3.3 | 2.1 (0.6-7.0) |
| A.D.5, A.D.5.1 and A.D.5.2 | 2.5 (0.6-10.3) |

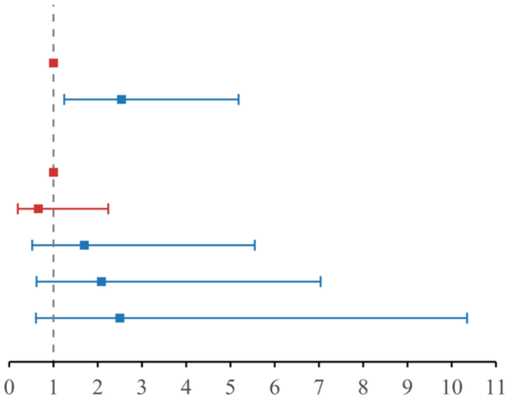


**Figure S3. Risk of nirsevimab immunization failure according to RSV groups and lineages without imputation (n=230).**

| Analysis | Cases | Control patients | Effectiveness (95% CI) |
|---|---|---|---|
| | no. who received nirsevimab/total no. (%) | | |
| **Overall effectiveness** | 354/2254 (15.7) | 414/737 (56.2) | 82.5 (78.2-85.9) |
| **Effectiveness according to RSV subtypes** | | | |
| RSV-A | 257/1531 (16.8) | 414/737 (56.2) | 79.4 (73.7-83.8) |
| RSV-B | 97/723 (13.4) | 414/737 (56.2) | 89.1 (85.3-91.9) |
| **Effectiveness according to RSV lineages** | | | |
| A.D.1, A.D.1.5 and A.D.1.6 | 114/815 (14.0) | 414/737 (56.2) | 84.8 (79.2-88.9) |
| A.D.3, A.D.3.1 and A.D.3.3 | 98/333 (29.4) | 414/737 (56.2) | 67.9 (56.0-76.6) |
| A.D.5, A.D.5.1 and A.D.5.2 | 45/383 (11.7) | 414/737 (56.2) | 79.5 (66.9-87.3) |
| B.D.4.1 and B.D.4.1.1 | 23/194 (11.9) | 414/737 (56.2) | 90.5 (84.5-94.2) |
| B.D.E.1, B.D.E.1.1 and B.D.E.1.2 | 74/529 (14.0) | 414/737 (56.2) | 88.3 (83.8-91.6) |

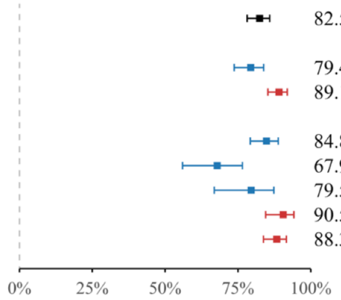


**Figure S4. Effectiveness of nirsevimab against hospitalization for RSV bronchiolitis in infants, excluding those were immunized less than 7 days before hospitalization (n=2991).**

| Analysis | Cases | Control patients | Effectiveness (95% CI) |
|---|---|---|---|
| | no. who received nirsevimab/total no. (%) | | |
| **Overall effectiveness** | 177/1413 (12.5) | 134/288 (46.5) | 79.8 (70.6-86.2) |
| **Effectiveness according to RSV subtypes** | | | |
| RSV-A | 168/1091 (15.4) | 134/288 (46.5) | 71.8 (58.3-80.9) |
| RSV-B | 9/322 (2.8) | 134/288 (46.5) | 97.1 (93.0-98.8) |
| **Effectiveness according to RSV lineages** | | | |
| A.D.1, A.D.1.5 and A.D.1.6 | 95/648 (14.7) | 134/288 (46.5) | 74.8 (61.0-83.7) |
| A.D.3, A.D.3.1 and A.D.3.3 | 28/88 (31.8) | 134/288 (46.5) | 47.4 (-17.9-76.5) |
| A.D.5, A.D.5.1 and A.D.5.2 | 45/355 (12.7) | 134/288 (46.5) | 74.2 (54.7-85.3) |
| B.D.4.1 and B.D.4.1.1 | 4/105 (3.8) | 134/288 (46.5) | 96.4 (87.8-98.9) |
| B.D.E.1, B.D.E.1.1 and B.D.E.1.2 | 5/217 (2.3) | 134/288 (46.5) | 97.5 (92.1-99.2) |

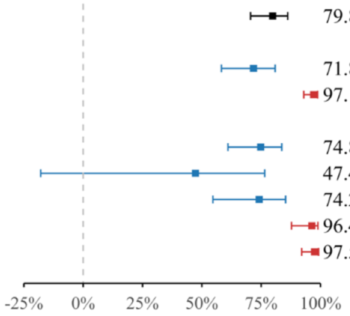


**Figure S5. Effectiveness of nirsevimab against hospitalization for RSV bronchiolitis in infants in 2023-2024 epidemic RSV season.**

| Analysis | Cases | Control patients | Effectiveness (95% CI) |
|---|---|---|---|
| | no. who received nirsevimab/total no. (%) | | |
| **Overall effectiveness** | 195/859 (22.7) | 289/458 (63.1) | 83.7 (78.6-87.7) |
| **Effectiveness according to RSV subtypes** | | | |
| RSV-A | 98/449 (21.8) | 289/458 (63.1) | 84.9 (79.0-89.1) |
| RSV-B | 97/410 (23.7) | 289/458 (63.1) | 83.5 (77.1-88.2) |
| **Effectiveness according to RSV lineages** | | | |
| A.D.1, A.D.1.5 and A.D.1.6 | 24/172 (14.0) | 289/458 (63.1) | 92.4 (87.3-95.5) |
| A.D.3, A.D.3.1 and A.D.3.3 | 74/249 (29.7) | 289/458 (63.1) | 75.7 (65.1-83.1) |
| A.D.5, A.D.5.1 and A.D.5.2 | 0/28 (0.0) | 289/458 (63.1) | 100 (CI unestimated) |
| B.D.4.1 and B.D.4.1.1 | 21/91 (23.1) | 289/458 (63.1) | 85.2 (74.3-91.5) |
| B.D.E.1, B.D.E.1.1 and B.D.E.1.2 | 76/319 (23.8) | 289/458 (63.1) | 83.2 (76.0-88.3) |

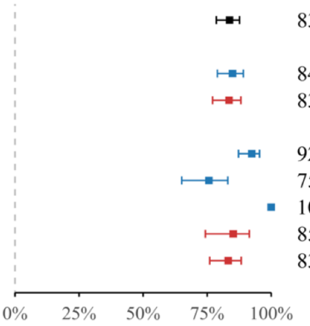


**Figure S6. Effectiveness of nirsevimab against hospitalization for RSV bronchiolitis in infants in 2024-2025 epidemic RSV season.**

| Analysis | Cases | Control patients | Effectiveness (95% CI) |
|---|---|---|---|
| | no. who received nirsevimab/total no. (%) | | |
| **Overall effectiveness** | 373/2273 (16.4) | 423/746 (56.7) | 85.0 (82.1-87.6) |
| **Effectiveness according to RSV subtypes** | | | |
| RSV-A | 267/1541 (17.3) | 423/746 (56.7) | 84.1 (80.6-86.9) |
| RSV-B | 106/732 (14.5) | 423/746 (56.7) | 87.1 (83.4-90.0) |
| **Effectiveness according to RSV lineages** | | | |
| A.D.1, A.D.1.5 and A.D.1.6 | 119/820 (14.5) | 423/746 (56.7) | 87.0 (83.5-89.9) |
| A.D.3, A.D.3.1 and A.D.3.3 | 102/337 (30.3) | 423/746 (56.7) | 66.9 (56.5-74.9) |
| A.D.5, A.D.5.1 and A.D.5.2 | 45/383 (11.7) | 423/746 (56.7) | 89.8 (85.8-92.9) |
| B.D.4.1 and B.D.4.1.1 | 25/196 (12.8) | 423/746 (56.7) | 88.8 (82.9-93.0) |
| B.D.E.1, B.D.E.1.1 and B.D.E.1.2 | 81/536 (15.1) | 423/746 (56.7) | 86.4 (82.1-89.8) |

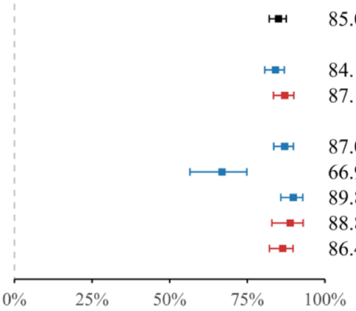


**Figure S7. Unadjusted effectiveness of nirsevimab against hospitalization for RSV bronchiolitis in infants (n=3019).**

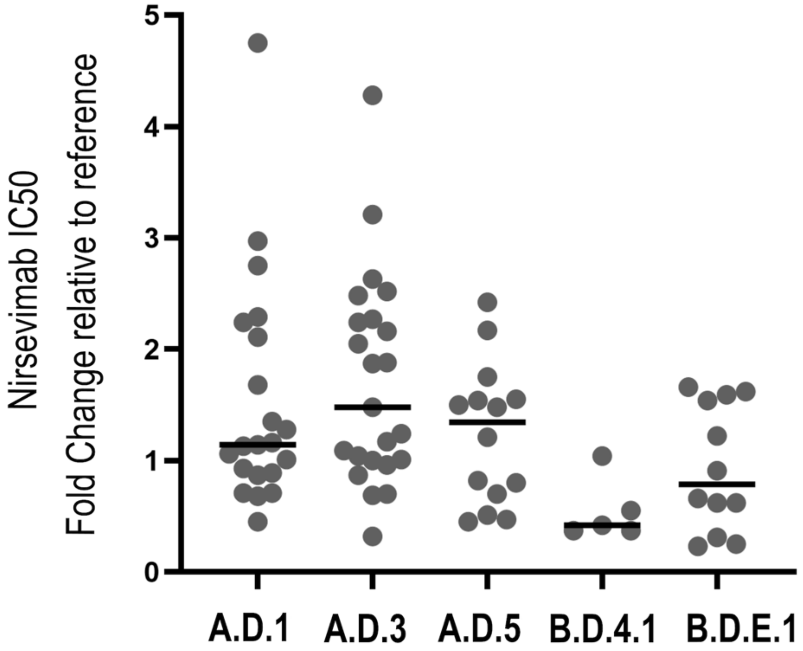


**Figure S8. Nirsevimab susceptibility of RSV isolates.**
IC50=half-maximal inhibitory concentration.
